# Non-neutralizing secretory IgA and T cells targeting SARS-CoV-2 spike protein are transferred to the breastmilk upon BNT162b2 vaccination

**DOI:** 10.1101/2021.05.03.21256416

**Authors:** Juliana Gonçalves, A. Margarida Juliano, Nádia Charepe, Marta Alenquer, Diogo Athayde, Filipe Ferreira, Margarida Archer, Maria João Amorim, Fátima Serrano, Helena Soares

## Abstract

In view of data scarcity to guide decision-making in breastfeeding women, we evaluated how mRNA vaccines impact immune response of lactating health care workers (HCW) and the effector profile of breast milk transferred immune protection. We show that upon BNT162b2 vaccination, immune transfer via milk to suckling infants occurs through secretory IgA (SIgA) and T cells. Functionally, spike-SIgA was non-neutralizing and its titers were unaffected by vaccine boosting, indicating that spike-SIgA is produced in a T-cell independent manner by mammary gland. Even though their milk was devoid of neutralizing antibodies, we found that lactating women had higher frequencies of RBD-reactive circulating memory B cells and more RBD-IgG antibodies, when compared to controls. Nonetheless, blood neutralization titers in lactating and non-lactating HCW were similar. Further studies are required to determine transferred antibodies and spike-T cells complete functional profile and whether they can mediate protection in the suckling infant.

**Highlights:**

- Milk and blood responses to BNT162b2 vaccine are initially isotype discordant
- Immune transfer via milk to suckling infants occurs by spike-reactive SIgA and T cells
- Spike-reactive SIgA in the breastmilk is non-neutralizing and T-cell independent
- Lactating vs non-lactating HCW had distinct cellular responses, despite similar NT50

## Introduction

Initial COVID-19 clinical trials of mRNA vaccines excluded lactating women, causing a dearth of data to guide vaccine decision-making by health authorities ^1^. Even after vaccine emergency authorization, breastfeeding HCW have been advised to discontinue breastfeeding upon receiving COVID-19 mRNA vaccine ^2^. This is especially worrisome since infants are the children’s age group most affected by COVID-19 ^3,4^. In view of the physiological alterations observed in lactating women and of the crucial role of breastmilk in providing immunity to the suckling infant, there is a pressing need to foresee how mRNA vaccines impact immune responses in lactating mothers and to uncover the effector profile of breast milk transferred immune protection.

Infants have an immature immune system and rely on the transfer of maternal immune cells and antibodies via the breastmilk to provide them with immunity ^5-9^. Breastmilk humoral and cellular content changes during lactation, mirroring the development of the infant’s own immune system and digestive tract, but is present in the milk during at least the first year of infant life ^6,10,11^.

Human breastmilk contains a wide variety of immunoglobulins, including IgA (∼90%), IgM (∼8%) and IgG (∼2%) ^12^. While human milk IgG mostly originates from the blood, milk IgA and IgM originate from mucosa-associated lymphatic tissue (MALT) ^13,14^. At the mucosa sites, IgA and IgM are secreted in the form of polymeric antibodies complexed to j-chain and secretory component proteins ^13^. The secretory component plays a critical role in protecting SIgA and IgM from proteolytic cleavage in the gut, facilitating their digestive traffic, systemic uptake and tissue distribution in the infant ^14^. Importantly, neutralizing SIgA antibodies elicited through maternal vaccination protect suckling infants from respiratory disease upon transfer via breastmilk ^15^. Few recent reports have documented IgA presence in the breastmilk in response to COVID-19 mRNA vaccines ^16-20^. Nonetheless, it remains to be addressed whether vaccine elicited milk IgA is produced in the mammary mucosa in its secretory SIgA form or if it is provided as monomeric IgA form by the blood. Moreover, it is currently unknown if vaccine elicited milk IgA can confer immune protection to the infant through viral neutralization.

In addition to antibodies, breast milk contains several types of maternal immune cells, including B and T cells ^10,21,22^. Milk lymphocytes are more activated and/or differentiated than their blood counterparts, with milk T cells comprising almost exclusively of effector memory cells and with B cells containing mainly class-switched IgD-memory B cells and plasma cells ^23-27^. Several lines of evidence support that milk B and T cells are capable of withstanding gastric environment ^28-30^, enter blood circulation ^31,32^ and be distributed into infant tissues ^33,34^. Recent studies including human-based ones have indicated that transfer of maternal lymphocytes via breast milk greatly assists the newborn’s immune system ^14^. It has been previously shown that mRNA vaccines induce spike reactive B and T cells in the blood ^35,36^. However, it remains to be addressed whether those vaccines can elicit local mucosal T and B cell responses which could be transferred to the suckling infant via breastmilk.

Prolactin, the hormone that promotes lactation, also functions as a cytokine stimulating lymphocyte proliferation and cytokine production ^37^. Consequently, prolactin drives a unique immune profile, composed by increased phagocytic and cytolytic activities and B and T cell activation. A couple of recent studies have shown that antibody production between lactating and non-lactating women receiving COVID-19 mRNA vaccines is similar ^17,18^. Nonetheless, how the mRNA vaccines impart on the cellular immune response of lactating women has so far remained unaddressed.

Here we sought to gauge the effects of mRNA vaccines on the humoral and cellular immune responses of breastfeeding women and to uncover breast milk effector immune composition.

## Results

### Milk and blood response to vaccine 1^st^ dose are isotype discordant and non-neutralizing

We collected paired breastmilk and blood samples from 14 lactating, a median of 10 days after first (interquartile range (IQR), 8-12 days) and second (IQR, 9-10 days) Pfizer BNT162b2 mRNA vaccine administration. Infection by SARS-CoV-2 had not been reported in any of the study participants. Demographic data of the study is contained in Tables S1 and S2.

We first looked at humoral response in the breastmilk and blood compartments ∼10 days post first vaccine dose, when protection conferred by BNT162b2 is starting ^38,39^, by assessing IgG, IgA and IgM antibodies to SARS-CoV-2 trimeric spike protein. All lactating women had anti-spike antibodies in circulation with 9/14 triple positive for IgG, IgA and IgM, 2/14 double positive for IgG and IgM, 2/14 single positive for IgG and 1/14 single positive for IgM (Fig, 1A; left graph, orange). In contrast, 8/14 presented anti-spike antibodies in the breastmilk, overwhelmingly of IgA isotype (8/8) with one donor having both IgG and IgA (Fig, 1A; right graph, purple). Anti-spike IgG and IgA antibody levels were higher in the blood (Fig. 1B; orange) and did not correlate with donors’ age (Fig. 1C).

**Figure 1.**
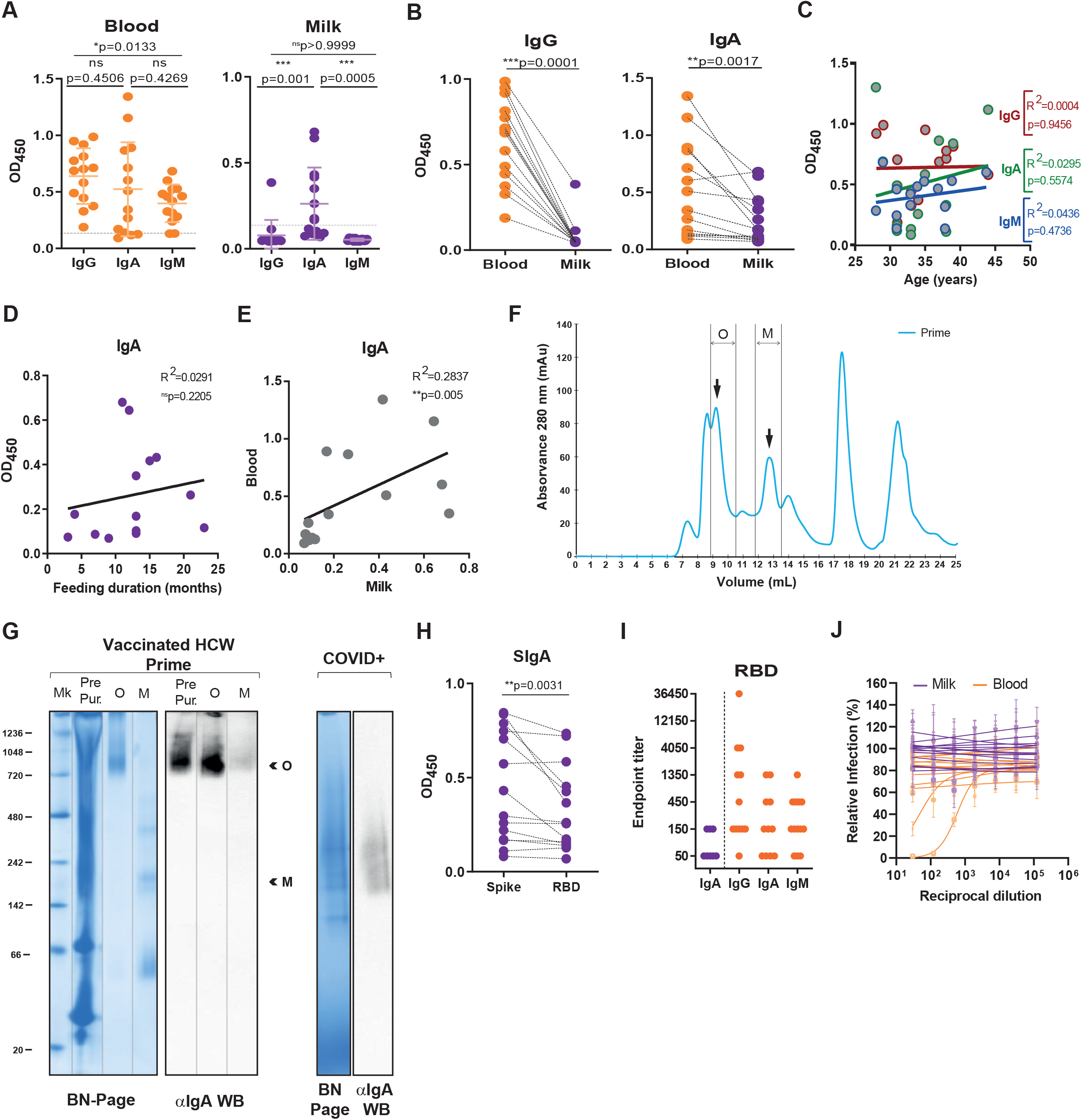
Early response to vaccine 1^st^ dose in the milk and in the blood are isotype discordant and non-neutralizing. (A) Levels of IgG, IgA and IgM against SARS-CoV-2 trimeric spike protein in the plasma (left graph; orange) and skim milk (right graph: purple) diluted at 1:50 in breastfeeding HCW (n=14) at ∼10 days post first dose of BNT162b2 mRNA vaccine, measured by absorbance at 450 nm (OD_450_). Dashed line indicates assay cut off. (B) Donor matched analysis of OD_450_ values for IgG and IgA in plasma (orange) and in skim milk (purple) diluted at 1:50. (C) Correlation between blood IgG, IgA and IgM levels and donor’s age in years. (D) Correlation between milk IgA recognizing trimeric spike protein after the first vaccine dose and feeding duration in months. (E) Correlation between trimeric spike reactive IgA in the plasma versus skim milk. (F) Illustrative size exclusion chromatogram of skim milk. Two fractions, designated O and M, suspected to correspond to oligomeric SIgA and monomeric immunoglobulins, were collected. (G) Unfractionated skim milk (Pre Pur) together with fractions O and M, suspected to correspond to oligomeric SIgA and monomeric immunoglobulins, and unfractionated serum sample from a COVID-19 patient (COVID+) were analysed through Blue Native PAGE (left) and α-IgA western blotting (right). (H) OD_450_ values of size exclusion chromatography purified SIgA recognizing trimeric spike protein or its RBD domain. (I) Endpoint titers for skim milk IgA (purple) and plasma IgG, IgA and IgM (orange) recognizing spike RBD domain. (J) Neutralization curves for plasma (orange) and skim milk (purple) for SARS-CoV-2 pseudotyped virus. p values determined by ANOVA, post-hoc Turkey’s and Friedman, post-hoc Dunn’s when comparing 3 groups. p values determined by parametric paired *t* test and by non-parametric paired Wilcoxon test when appropriate. Pearson correlation was used for parametric data and Spearman correlation for non-parametric data. ***p<0.001, **p<0.01, *p<0.05, ns = not significant.

Lactation stage affects breastmilk immune composition. Conversion from colostrum (until 3-5 days after birth) to transitional milk (from day 3-5 to 2 weeks postpartum) and finally to mature milk (from week 2 to the end of lactation) is accompanied by a decrease in milk immune components. Nonetheless, mature milk immune contents remain constant through the duration of lactation period ^14^. Accordingly, in our cohort, composed exclusively by mature breastmilk samples, milk IgA levels did not vary by breastfeeding duration (Fig. 1D). Interestingly, we detected a trend between milk and blood anti-spike IgA levels (Fig. 1E). Circulating and mucosal IgA are molecularly distinct. Whereas SIgA originated in the mammary mucosa is secreted in its dimeric, trimeric or tetrameric forms, blood IgA occurs as monomers ^40^. As mRNA vaccines are better suited at inducing systemic rather than mucosal immunity, we sought to identify the source of IgA in the breast milk. To this end, we fractionated skim milk through size exclusion chromatography and collected all fractions. Based on the calculated molecular weight (MW) range for each peak (Fig. S1) and previous protein profiles of human milk ^41^, oligomeric forms of SIgA (MW: 400-800 kDa) and monomeric immunoglobulins (MW: 90-170 kDa) were expected to elute in peaks O and M, respectively (Fig. 1F). We confirmed this by first running fractions O and M together with non-fractioned milk on BN-PAGE (Fig. 1G). The protein profile of non-fractionated milk is consistent with the protein profile observed on the size exclusion chromatogram (Fig. 1 F, G). Fraction O shows a smeared band above 720 kDa, likely corresponding to SIgA tetramers. Fraction M depicts a single band at ∼150 kDa, probably consisting of monomeric immunoglobulins. The western blotting against human IgA corroborates the presence of IgAs in the pre-purified sample and in fraction O, but not in fraction M (Fig. 1G). These data exclude the possibility that milk spike-reactive IgA originates from the blood and indicate that the ∼150 kDa band visible in the native gel likely corresponds to IgG (Fig. 1G). Finally, we verified that SIgA recognized both trimeric spike protein and its RBD domain (Fig. 1H), confirming that anti-spike IgA antibodies are produced by the mammary mucosa in the form of SIgA and not sourced from the blood. Curiously, SIgA displayed higher reactivity towards trimeric spike than towards its RBD domain, raising questions about its neutralization activity.

To gain insight into the possible neutralizing properties of milk and blood antibodies, we first assessed their endpoint titers for the RBD domain ^42^. Milk anti-RBD SIgA endpoint titers were lower when compared to blood IgA, IgG and IgM (Fig. 1I). We next determined their virus-neutralizing activity through a microneutralization assay using spike pseudotyped lentivirus infection of ACE-2 receptor expressing 293T cell line ^43^. At 10d post vaccine prime, milk IgA had no neutralizing activity (Fig. 1J; purple). Curiously, even though several blood samples had moderate (1:450) and high (>1:1350) endpoint titers for IgG, IgA and IgM, only one blood sample (1/14) displayed a weak neutralizing activity (Fig. 1 I, J).

Altogether our data shows that SIgA is produced early (d10) by the mammary mucosa in response to the first dose of BNT162b2 vaccine. This early mammary production of SIgA likely accounts for the distinct immunoglobulin composition between the milk and blood compartments. Regardless, neither blood nor milk antibodies possess neutralizing capacity at this point.

### SIgA in breastmilk remains non-neutralizing even after vaccine second dose

While, circulating neutralizing IgG to BNT162b2 vaccine are optimally detected after boost ^44^, it is currently unknown whether vaccine induced milk antibodies will provide neutralizing protection to the sucking infant ^16-18^. Contemporaneously with IgG surge in the blood (Fig. 2 A, B; orange) at a median of 10 days after vaccine second dose, we detected anti-spike and anti-RBD IgG (14/14), but not IgM (0/14), in the milk (Fig. 2 A, B; purple). Interestingly, the frequency of milk samples containing spike- and RBD-reactive SIgA was not altered (8/14), their milk levels remained constant upon vaccine boost and did not correlate with feeding duration (Fig. 2 A-C; purple). Similarly, circulating levels of IgA recognizing spike protein remained unaltered by vaccine boost (Fig. 2A; orange). However, circulating anti-RBD IgA levels were augmented after vaccine second dose (Fig. 2B; orange).

**Figure 2.**
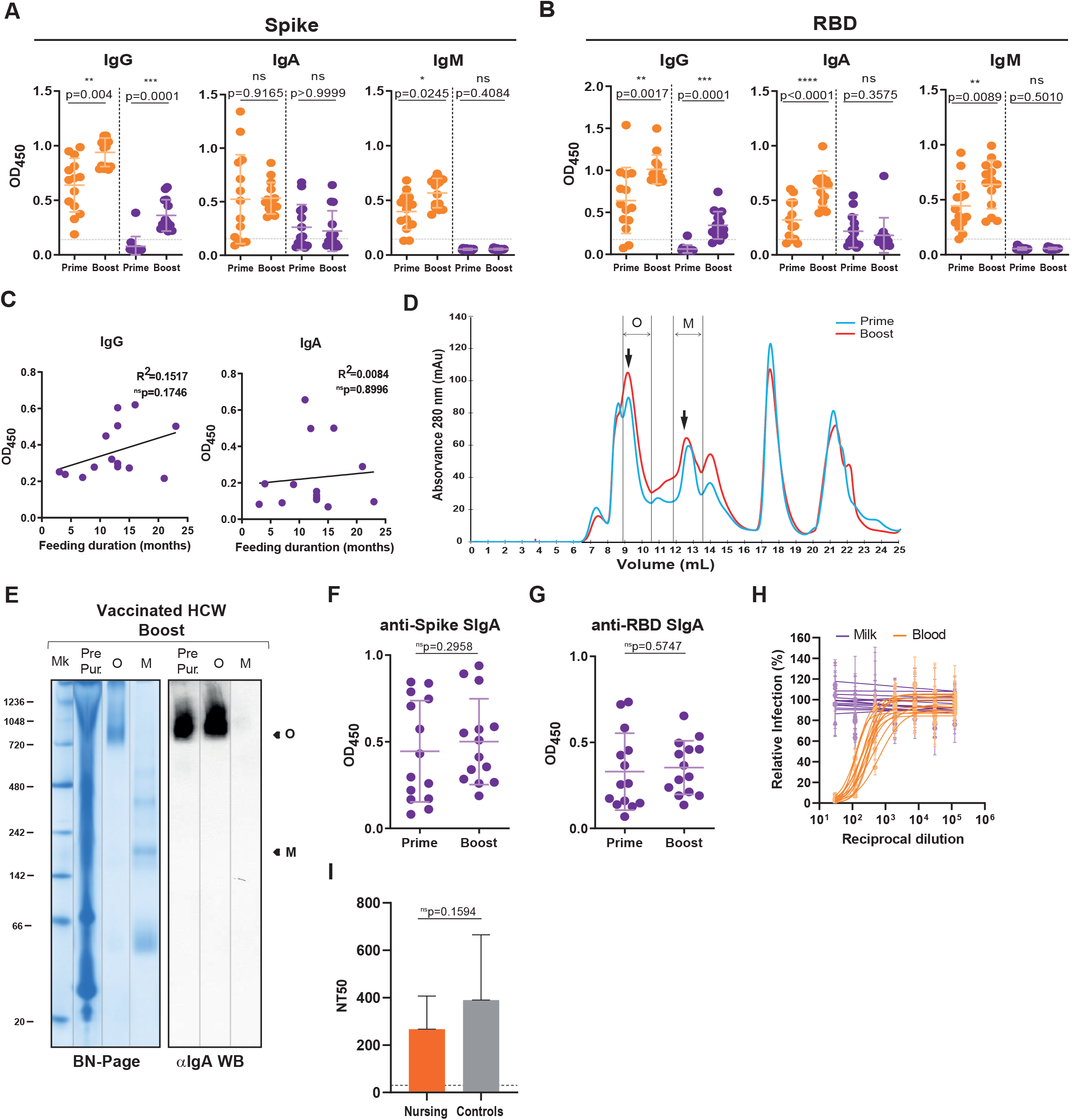
Neutralizing antibodies are found in the blood but not in the milk after vaccine second dose. (A) Comparison of IgG, IgA and IgM, recognizing trimeric spike protein, at median of 10 days after the first (prime) and median of 10 days after the second (boost) of BNT162b2 vaccine in the plasma (orange) and in skim milk (purple) diluted at 1:50, measured by absorbance at 450 nm (OD_450_). Dashed line indicates test cut off. (B) Comparison of IgG, IgA and IgM, recognizing spike RBD domain, performed as in (A). Dashed line indicates test cut off. (C) Correlation between milk IgG (left) and IgA (right) recognizing trimeric spike protein and feeding duration in months. (D) Illustrative size exclusion chromatogram of a skim milk samples from the second (red) and first (blue) doses. Two fractions, designated O and M, suspected to correspond to oligomeric SIgA and monomeric immunoglobulins, were collected. (E) Unfractionated skim milk (Pre Pur) together with fractions O and M, suspected to correspond to oligomeric SIgA, respectively, were analysed through Blue Native PAGE (left) and α-IgA western blotting (right). (F, G) Comparison between the OD_450_ values of SEC-purified SIgA recognizing trimeric spike protein (F) or its RBD domain (G) after the first (prime) and second (boost) vaccine doses. (H) Neutralization curves for plasma (orange) and skim milk (purple) for SARS-CoV-2 pseudotyped virus. (I) Neutralization titers (NT50) for SARS-CoV-2 pseudotyped virus in the plasma of nursing (orange) and non-breastfeeding (control; grey) HCW. p values determined by parametric paired *t* test and by non-parametric paired Wilcoxon test when appropriate. For unpaired data, Man-Whitney test was used when appropriate. Spearman correlation was used for correlative analysis. ****p<0.0001, ***p<0.001, **p<0.01, *p<0.05, ns = not significant.

Next, we wanted to know whether vaccine boost altered SIgA profile in the milk. As before, we fractionated skim milk through size exclusion chromatography and ascertained the composition of fractions O and M by western blot against human IgA (Fig. 2D, E). As can be observed in the overlay of a paired sample, the size exclusion chromatograms yield a similar profile after vaccine 1^st^ and 2^nd^ doses (Fig. 2D), which was similarly observed for all the breastmilk samples (Fig. S2). We compared SIgA reactivity to trimeric spike and its RBD domain after the vaccination regimen by ELISA and found that there were no differences between vaccine the first and second vaccine doses (Fig. 2 F, G). The fact that vaccine boost has no effect in the milk levels of spike- and RBD-reactive SIgA, suggests that SIgA production by the mammary mucosa in response to the vaccine is made in a T cell-independent manner.

As expected, all the plasmas from fully vaccinated lactating HCW were neutralizing (NT50: 231.24; IQR, 189.58–298.05) and comparable to an aged-matched female HCW cohort (NT50: 315.54; IQR, 212.31–456.94) (Fig. 2 H, I) and to previous reports for BNT162b2 vaccine ^36,44,45^. Importantly, regardless of the increase in anti-spike SIgA tetramerization status none of the milk samples displayed neutralizing activity, supporting the view that SIgA tetramerization might be implicated in increasing target breadth rather than neutralization activity^40^.

In all, our data suggest that milk vaccine induced SIgA is likely produced in a T cell independent manner. Moreover, spike- and RBD-reactive SIgA transferred by the breast milk is unlikely to provide protection to the suckling infant through SARS-CoV-2 neutralization.

### Lactating HCW have higher frequency of RBD-reactive memory B cells and RBD-recognizing antibodies in circulation than controls

While human IgA secreting B cells are retained in the mammary gland, functional IgG secreting B cells can be transferred to the suckling infant via milk ^14,25,46^. We sought to evaluate whether RBD-reactive B cells were present in the milk of breastfeeding HCW after both the first and second vaccine doses. We were only able to detect CD3^-^CD19^+^ B cells in 5 out of 14 milk samples (Fig 3 A, B). Consistent with previous report ^25^, milk B cells were overwhelming IgD^-^ illustrating that they had undergone class-switch (Fig 3A). Due the limited number of B cells detected we were not able to assess the presence of RBD-reactive milk B cells. Mainly due to hormonal changes, lactating women display distinct immune responses ^37^. To fully evaluate the B cell immune response to BTN162b2 vaccine in this population, we determined the frequency of antibody secreting RBD-reactive plasmablasts and memory B cells (Fig 3 B, C). A clear population of RBD-binding IgD^-^ B cell population could be detected at ∼10 days post vaccine first dose (Fig 3 B, C; prime), whose frequency (∼1.7%) remained unaltered upon vaccine boost (Fig 3 B, C; boost). We then analyzed the frequency of RBD-reactive plasmablasts and memory B cells. Both RBD-reactive plasmablasts and memory B cells were detectable after vaccine prime (Fig. 3 B, C). Upon vaccine boost, only plasmablasts appear to increase in frequency (Fig. 3 B, C). Curiously, when compared to age- and gender-matched controls, in breastfeeding HCW the effector B cell response to RBD was skewed toward memory B cells with concomitant decrease in short-lived plasmablasts (Fig. 3C). Memory B cells have been proposed to play a key role in mounting recall responses to COVID-19 mRNA vaccines ^44^. Accordingly, breastfeeding HCW registered higher titers of circulating RBD-reactive IgG and IgA antibodies than controls (Fig. 3D). Finally, we found no correlation between the frequency of RBD-reactive total B cells or RBD-reactive memory B cells and spike/RBD-reactive IgG levels or with neutralization titers (Fig. 3 E, F).

**Figure 3.**
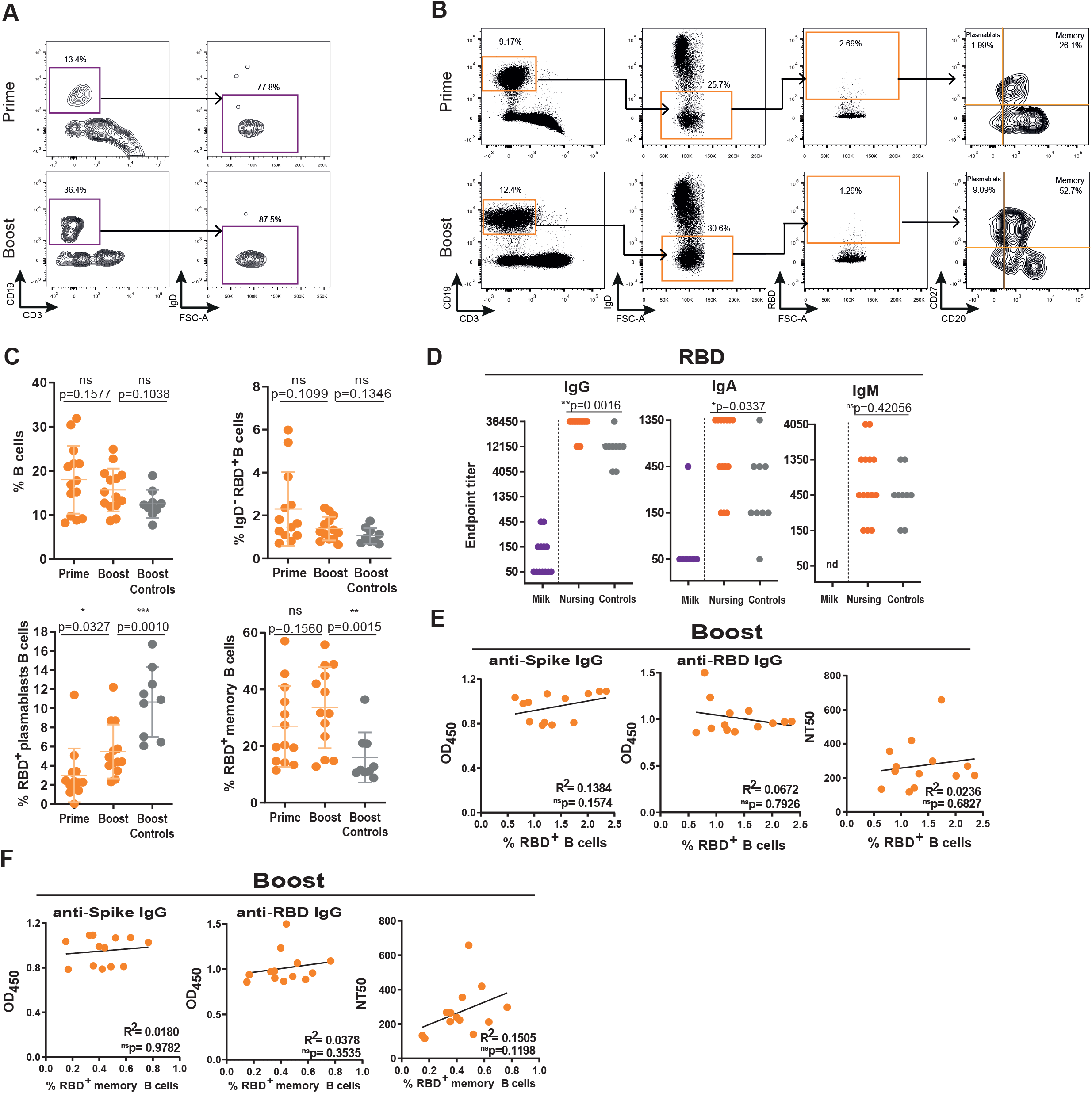
Lactating HCW have higher frequency of RBD-reactive memory B cells and RBD-recognizing antibodies in circulation than controls. (A) Gating strategy for CD3^-^CD19^+^IgD^-^ in the milk, after first (prime) and second (boost) doses of BNT162b2 vaccine in breastfeeding HCW. (B) Gating strategy for circulating RBD-reactive CD3^-^CD19^+^IgD^-^CD20^-^CD27^+^ plasmablasts and CD3^-^CD19^+^IgD^-^CD20^+^CD27^+^ memory B cells. (C) Cumulative frequency of circulating CD3^-^CD19^+^ (top left) total B cells, RBD reactive CD3^-^CD19^+^IgD^-^ B cells (top right), CD3^-^CD19^+^IgD^-^CD20^-^CD27^+^ plasmablasts (bottom left) and CD3^-^CD19^+^IgD^-^CD20^+^CD27^+^ memory B cells (bottom right) after first (prime) and second (boost) vaccine doses for nursing (orange) and after the second (boost) vaccine dose for non-breastfeeding (control; grey). (D) Endpoint titers for RBD-specific IgG, IgA and IgM from skim milk (purple) and plasma (orange) of nursing and from plasma (grey) of non-breastfeeding (control; grey) HCW. nd = non-detectable (E) Correlation between spike-specific IgG (left), RBD-specific IgG (middle), and neutralization titers (right; NT50) and the frequency of RBD-reactive CD3^-^ CD19^+^IgD^-^ B cells, upon vaccine boosting. (F) Correlation between spike-specific IgG (left), RBD-specific IgG (middle), and neutralization titers (right; NT50) and the frequency of RBD-reactive CD3^-^ CD19^+^IgD^-^CD20^+^CD27^+^ memory B cells, upon vaccine boosting. p values determined by parametric paired *t* test and by non-parametric paired Wilcoxon test when appropriate. For unpaired data, *t* test was used for parametric data and Man-Whitney test was used for non-parametric data when appropriate. Spearman correlation was used for correlative analysis. ***p<0.001, **p<0.01, * p<0.05, ns = not significant.

Although we were not able to detect RBD-reactive B cells, we found that lactating HCW had higher frequencies of RBD-reactive circulating memory B cells and higher RBD-IgG antibodies, when compared to controls.

### Spike-specific T cells are transferred through breastmilk after vaccine boost

Emerging evidence suggests the requirement of both antibody-mediated and T cell-mediated immunity for effective protection against SARS-CoV-2 ^47,48^. Importantly, these mammary gland T cells are transferred through the milk to the nursing infant and mediate immune protection ^27,49^.

To detect trimeric spike specific CD4^+^ T cells in the milk and in circulation, we used an Activation Induced Marker (AIM) assay using OX40 and CD25 dual expression to detect spike reactivity ^50-52^. Out of 14 breastfeeding HCW, we could only robustly detect CD4^+^ T cells in the milk of 5 donors. After vaccine second dose, spike-reactive OX40^+^CD25^+^CD4^+^ T cells could be detected in 100% of milk samples with detectable CD4^+^ T cells (Fig. 4 A, B). Suggesting that in addition to non-neutralizing antibodies, spike-reactive CD4^+^ T cells are transferred through the breastmilk, upon mRNA vaccination.

**Figure 4.**
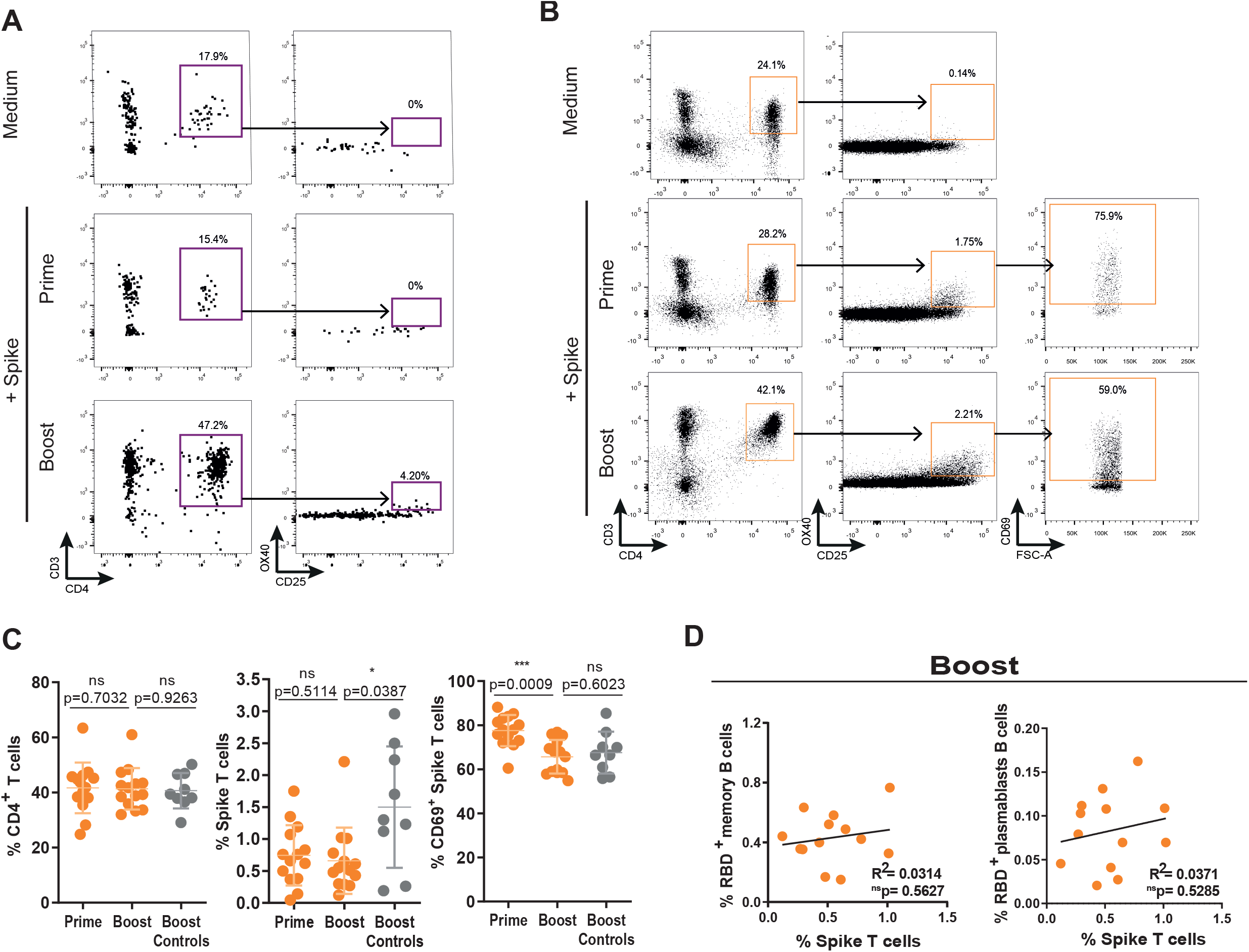
Spike-specific T cells are transferred through breastmilk after vaccine boost. (A) Gating strategy for CD3^+^CD4^+^OX40^+^CD25^+^ spike specific T cells in the milk, after first (prime) and second (boost) doses of BNT162b2 vaccine in breastfeeding HCW. (B) Gating strategy of circulating CD3^+^CD4^+^OX40^+^CD25^+^ spike specific T cells, after first (prime) and second (boost) doses of BNT162b2 vaccine in breastfeeding HCW. (C) Cumulative frequency of CD3^+^CD4^+^ (left), CD3^+^CD4^+^OX40^+^CD25^+^ (middle) and of CD3^+^CD4^+^OX40^+^CD25^+^CD69^+^ (right) circulating T cells after first (prime) and second (boost) vaccine doses for nursing (orange) and after the second (boost) vaccine dose for non-breastfeeding (control; grey). (D) Correlation between the frequency of RBD-reactive CD3^-^CD19^+^IgD^-^ CD20^+^CD27^+^ memory B cells (right) and RBD-reactive CD3^-^CD19^+^IgD^-^CD20^-^ CD27^+^ plasmablasts cells (left) with the frequency of CD3^+^CD4^+^OX40^+^CD25^+^ circulating T cells after second vaccine dose. p values determined by parametric paired *t* test and by non-parametric paired Wilcoxon test when appropriate. For unpaired data, *t* test was used for parametric data and Man-Whitney test was used for non-parametric data when appropriate. Pearson correlation was used for correlative analysis. ***p<0.001, *p<0.05, ns = not significant.

All breastfeeding vaccinated HCW possessed spike reactive OX40^+^CD25^+^CD4^+^ T cells (median, 0.71%; IQR, 0.38–1.07) in circulation right after vaccine prime, and their frequency was not altered by subsequent boost (Fig 4 B, C), even though their activation state measured by CD69 expression decreased with vaccine boost (Fig. 4 B, C). Curiously, it appears that after vaccine boost breastfeeding HCW tended to have less spike-reactive CD4^+^ T cells when compared to vaccinated controls (Fig. 4C). In view of the role of CD4^+^ T cells in B cell effector differentiation, we looked if there was an association between circulating spike-reactive T cells and RBD-reactive plasmablasts and memory B cells. There was no correlation between spike-reactive T cells and RBD-reactive plasmablasts or memory B cells (Fig. 4D).

All together these data show that in addition to non-neutralizing antibodies, milk also transfers spike-reactive T cells. The fact that spike-reactive T cells are only detectable in the milk after vaccine boost, reinforces the view that spike/RBD-sIgA, detected in the milk as early as 10 days post vaccine first dose, are likely the result of T-cell independent production.

## Discussion

Recent data from Brazil has brought to the forefront that within the pediatric population, infants are among the most susceptible to and present the highest COVID-19 fatality rate ^3,4,53^. Yet, lactating women were excluded from initial vaccine trials and lactating HCW were often advised by national health authorities to discontinue breastfeeding upon receiving COVID-19 mRNA vaccine ^2^. Evolved to provide immunity to the suckling infant, lactating women display a unique immune activation state. How mRNA vaccines impact this distinct immune state and which is the breadth and effector profile of milk transferred immune response remains poorly understood. Through a combination of serology, virus neutralization assays and size exclusion chromatography we identified, to the best of our knowledge for the first time, the secretory and neutralizing properties of breastmilk transferred antibodies and cellular immunity following BNT162b2 vaccination. Moreover, we immunophenotyped the B and T cell responses to the vaccine in the milk and blood of lactating women. All together, these findings indicate that a possible immune protection conferred by breastmilk will occur in a neutralization-independent T cell-dependent fashion. In addition, the higher frequency in RBD-memory B cells in lactating women raise the possibility of increased duration of vaccine conferred immune protection.

A dramatic plunge of COVID-19 cases starts as early as 12 days after BNT162b2 first vaccine dose ^38,39^. We found that early vaccine immune response in the blood and in the breastmilk was isotype discordant. Whereas, breastmilk contained exclusively low titers of spike-reactive IgA, circulating anti-spike antibodies spanned from low to high titers and through IgG, IgA, and IgM isotypes. These results highlight the functional compartmentalization between mucosal and the systemic immunity and the limitations of systemic administered BNT162b2 vaccine in eliciting a strong mucosal response ^54^. The function of antibodies produced within the mammary gland MALT is to provide protective immunity to the suckling infant through the secretion of polymeric antibodies complexed to j-chain and secretory component proteins ^13^. The secretory component is essential to ensure that milk antibodies are effectively transferred via the breastmilk by providing protection from proteolytic cleavage and by facilitating systemic uptake and tissue distribution in the infant ^13^. Spike-reactive SIgA were detected in the breast milk of COVID-19 patients ^55^, nonetheless whether SIgA was similarly present in the breast milk upon mRNA vaccination had remained unaddressed. By combining size exclusion chromatography with customized spike-ELISA, we concluded that vaccination does in fact originate spike-reactive SIgA, which is secreted predominately as tetramers. We detected SIgA in 57% of milk samples. Our detected milk SIgA prevalence is on the lower-end of the interval of milk IgA prevalence (61.8% to >80%) detected by others ^16-20^. This discrepancy can probably be ascribed to different experimental procedures. While we detected SIgA in milk samples that had been diluted 50-fold, these previous works either used undiluted milk samples or diluted them at a maximum of 5-fold.

Upon administration of influenza vaccine to pregnant women, neutralizing influenza-specific IgA are effectively transferred to breast milk ^15^. Whether mRNA vaccines similarly promote the transfer of neutralizing spike-specific IgA to breastmilk has remained unexplored. Our data elucidate that upon vaccination of lactating women with BNT162b2, milk secreted spike-reactive SIgA is non-neutralizing. This is in great contrast with COVID-19 infection, where neutralizing IgA occurs in the milk of infected lactating women ^56^. This is likely due to the fact that SARS-CoV-2 infection effectively primes airway and gut mucosa immunity, while the vaccine is defective in providing mucosal immunity. In fact, mucosal IgA secretion has been shown to play an important role in the early control of SARS-CoV-2 infection ^57^ and a recent paper has shown that SIgA dimers cloned from COVID-19 patients display a 7-fold higher neutralization capacity when compared to IgG ^58^. Importantly, our results show that spike-reactive SIgA titers are not altered by vaccine second dose and that its presence in the milk precedes the detection of spike-T cells, strongly suggesting that spike-reactive SIgA is produced in a T cell-independent manner. T cell-independent IgA responses result in the production of low affinity and polyreactive antibodies ^59^, which could help explain the absence of neutralizing function. Moreover, BNT162b2 induced circulating IgA has been proposed to be produced also in a T-cell independent manner ^60^. Nevertheless, milk transferred spike-reactive SIgA could be conferring protection through non-neutralizing actions, namely through Fc-related functions such as antibody dependent cytotoxicity, complement activation and phagocytosis.

An important and until now completely unexplored route for milk transferred immunity is through T cells. Previous studies have shown that milk transferred lymphocytes can survive the adverse environment of the digestive tract and seed in the infant’s tissues ^30-36^. This is due to a combination of a biochemical reaction between infant’s saliva and breast milk that protects lymphocytes from acid injury ^5-7^, a decrease in enzyme and acid content in the infant’s digestive tract ^61,62^, and an increase in gut permeability ^31^. Despite the strong experimental support that mRNA vaccine induced CD4^+^ T cells might play an important role in mediating protection ^45,63,64^ especially in suboptimal neutralizing antibodies settings ^65^, this is, to the best of our knowledge, the first report to detect spike-reactive T cells in the breast milk of vaccinated mothers. It is possible that milk transferred spike-reactive T cells might mediate protective function by seeding in the infant’s upper respiratory tract and gut. We found that spike-reactive T cells accounted for ∼3.5% of milk CD4^+^ T cells. Even though this frequency might seem low, it is worth to have in mind that a suckling infant ingest large volumes of milk daily, which implies the ingestion of significant quantities of spike-reactive T cells.

Prolactin, the hormone that induces milk production also functions as a cytokine that plays an important role in immune activation ^37^. Since breastfeeding women were excluded from initial vaccine trials, we sought to compare their response to BNT162b2 vaccine. We detected higher RBD-reactive IgG and IgA titers in lactating women, nonetheless their neutralization titers were undistinguishable from controls. These results are consistent with previous findings ^17,18^. In addition, we found that lactating women had higher frequencies of RBD-reactive memory B cells but lower frequencies of plasmablasts in circulation. Suggesting that the immune response to the vaccine in lactating women might be veered toward longer lasting memory in detriment of short-lived antibody production. Further studies will be needed to determine if these cellular differences are maintained in medium term and whether they will impact long term protection.

Here we show that upon BNT162b2 vaccination immune transfer to the breastmilk does not involve neutralizing antibodies. In fact, milk transferred T cells together with non-neutralizing antibodies might play an important role in mediating protection to SARS-CoV-2 infection in the infant. Our data also support the view that lactating women immune responses to the vaccine are qualitatively different and veered toward long-lasting memory B cell response, despite displaying equivalent neutralization antibody titers. It is worth mentioning that the results obtained represent only a snapshot of what is likely a dynamic immune response, using a small cohort. Future works using a larger sample size and longitudinal design are needed to determine whether the non-neutralizing antibodies and T cells contained in the breastmilk following BNT162b2 vaccination transfer immunity to the suckling infant and to elucidate their effector mechanisms.

## Data Availability

All data will be made available upon request

## Supplemental Figure Legends

**Figure S1.**
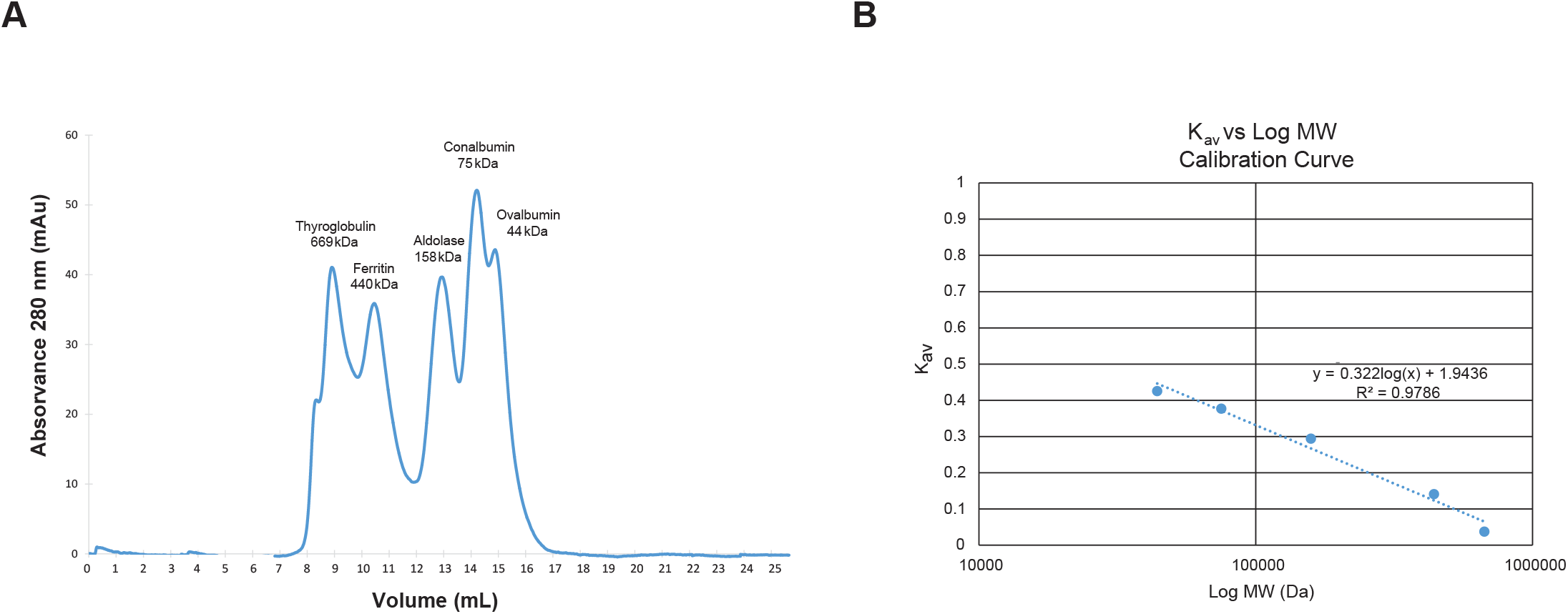
Standardization of molecular weight range of the chromatographic separation. **(A)** Chromatogram of all standard proteins run in a Superdex 200 increase 10/300 GL. HWM Filtration Calibration Kit (Cytiva) were used with the following proteins: thyroglobulin (669 kDa); ferritin (450 kDa), aldolase (158 kDa), conalbumin (75 kDa) and ovalbumin (44 kDa). These standard proteins were dissolved in bi-distilled water and their chromatographic profiles were obtained using an UV detector. **(B)** Graphical representation of calibration curve of partition coefficient (Kav) of each protein versus their respective molecular weight in Daltons. The Kav was calculated through the following formula: Kav=(Ve–V0)/(Vc-V0) where Ve is the elution volume of the protein, V0 is the void volume and Vc is the column bed volume. A dispersion graph Kav vs logMW was constructed. The equation obtained for the calibration curve is: Kav = -0.322log(MW) + 1.9436, where Kav is the partition coefficient and MW is the protein molecular weight (Da).

**Figure S2.**
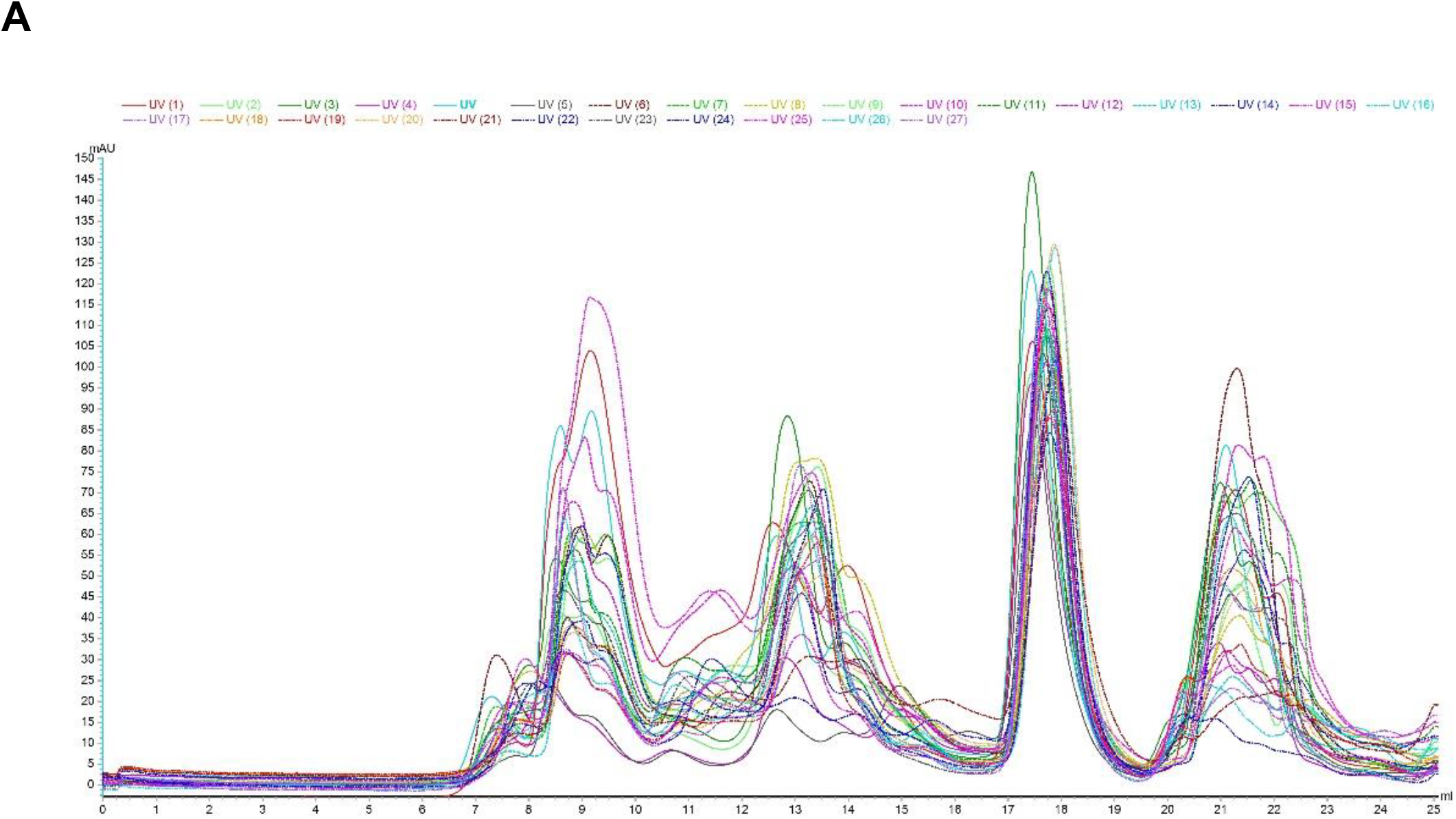
Size-exclusion chromatograms of breastmilk samples (Superdex 200 Increase 10/300 GL). **(A)** SEC-chromatograms of all breastmilk samples from first and second vaccine doses superposed.

## Acknowledgements

We are very grateful to all the participants of the study. We thank Cláudia Andrade at CEDOC Flow Cytometry platform for technical support. This work was supported by ESCMID and by Gilead Génese (PGG/009/2017) grants to HS, RESEARCH4COVID 19 (Ref 580) to MJA, EU H2020 projects No. 823780 and 871037 (iNEXT-Discovery) to MA, and by Fundação para a Ciência e Tecnologia (FCT) through Project MOSTMICRO-ITQB with refs UIDB/04612/2020 and UIDP/04612/2020 to MA and DA. JG, DA, MJA HS are supported by Fundação para a Ciência e Tecnologia (FCT) through PD/BD/128343/2017, BD/147987/2019, CEECIND/02373/2020 and CEECIND/01049/2020, respectively. Graphical abstract was created with BioRender.com.

## Author contributions

JG, AMJ, MA, DA, FF designed and performed experiments and analyzed the data. NC and FS enrolled the subjects and collected clinical data. MJA and MA provided critical expertise and insights. HS conceptualized the study, designed experiments, analyzed the data, supervised the project and wrote the manuscript. All authors discussed the results and commented on the manuscript.

## Competing interests

The authors declare no competing interests.

## STAR METHODS

### Study participants and human samples

Blood and breastmilk from 14 nursing mothers were collected between day 7 and 16 and between day 7 and 13 post-immunization with Pfizer BTN162b2 mRNA vaccine (Table S1). Blood from controls was collected between day 10 and 16 after vaccine second dose (Table S2). Blood was collected by venipuncture in EDTA tubes and breastmilk was collected with breast pump into sterile containers. Both biospecimens were immediately processed. All participants provided informed consent and all procedures were approved by NOVA Medical School ethics committee, in accordance with the provisions of the Declaration of Helsinki and the Good Clinical Practice guidelines of the International Conference on Harmonization.

### Peripheral blood and breastmilk cell isolation

Peripheral blood mononuclear cells (PBMCs) were isolated by density gradient centrifugation (Biocoll, Merck Millipore). Breast milk cells were isolated by centrifugation. Plasma and skim milk were respectively stored at -80°C or -20°C until further analysis. PBMCs and breastmilk mononuclear cells were suspended in freezing media (10% DMSO in FBS) and stored at -80°C until subsequent analysis.

### Flow cytometry for detection of SARS-CoV-2 reactive B and T cells

RBD was labelled with an available commercial kit according to manufactor’s instructions (life technologies, A20181). For detection of SARS-CoV-2 reactive T cells, cryopreserved PBMCs were rested for 1h at 37°C and then stimulated overnight with either 1 mg/mL of spike protein plus 5 μg/mL of anti-CD28 (CD28.2) (BioLegend) cross-linked with 2.5 μg/mL of anti-mouse IgG1 (RMG1-1) (BioLegend) or with medium alone (negative control). PBMCs were stained with a fixable viability dye eFluor™ 506 (invitrogen) and surface labelled with the following antibodies all from BioLegend: anti-CD3 (UCHT1), anti-CD4 (SK3), anti-OX40 (Ber-ACT35), anti-CD25 (M-A251), anti-CD69 (FN50), anti-CXCR-5 (J252D4), anti-CCR6 (G034E3), anti-CD19 (SJ25C1), anti-IgD (IA6-2), anti-CD27 (O323) and anti-CD20 (2H7) and also with the RBD labelling as described above. Cells were washed, fixed with 1% PFA and acquired in BD FACS Aria III equipment (BD Biosciences) and analysed with FlowJo v10.7.3 software (Tree Star).

### ELISA

Antibody binding to SARS-CoV-2 trimeric spike protein or its RBD domain was assessed by a previously described in-house ELISA assay ^42^ based on the protocol by Stadlbauer et al ^66^. Briefly, 96-well plates (Nunc) were coated overnight at 4°C with 0.5 μg/ml of trimeric spike or RBD. After blocking with 3% BSA diluted in 0.05% PBS-T, serially diluted plasma samples were added and incubated for 1 h at room temperature. Plates were washed and in incubated for 30 min at room temperature with 1:25,000 dilution of HRP-conjugated anti-human IgA, IgG and IgM antibodies (Abcam, ab97225/ab97215/ab97205) goat anti-human IgA/IgG/IgM-HRP secondary antibodies (diluted at in 1% BSA-0.05% PBS-T. Plates were washed and incubated with TMB substrate (BioLegend), stopped by adding phosphoric acid (Sigma) and read at 450nm. Endpoint titters were defined as the last dilution before the absorbance dropped below OD_450_ of 0.15. This value was established using plasma from pre-pandemic samples collected from subjects not exposed to SARS-CoV-2 ^42^. For samples that exceeded an OD_450_ of 0.15 at last dilution (1:10,9350), end-point titter was determined by interpolation ^67^. As previously described ^42^, in each assay we used 6 internal calibrators from 2 high-, 2 medium- and 2 low-antibody producers that has been diagnosed for COVID-19 through RT-PCR of nasopharyngeal and/or oropharyngeal swabs. As negative controls, we used pre-pandemic plasma samples collected prior to July 2019.

### Sample purification by size exclusion chromatography (SEC)

Skim milk samples were centrifuged at 20,000x *g*, for 5 min at 4°C, to remove precipitates or debris prior to SEC. A volume of 200 µL of each clarified sample was injected on a Superose 200 increase 10/300 GL column coupled to an AKTA pure FPLC system (Cytiva) with UV/Visible detector at 280 nm. PBS at pH 7.4 was used as elution buffer with a flow rate of 0.5 mL/min. A similar procedure was applied to a sample of blood serum from a COVID-19 positive patient to serve as control. Fractions of 0.5 mL were collected and gathered according to the peaks identified in the chromatogram to be analysed on blue native polyacrylamide gel electrophoresis (BN-PAGE) on NativePAGE 4–16% Bis-Tris gels (ThermoFisher Scientific) with NativeMark (ThermoFisher Scientific) as molecular weight marker and stained with ProBlue Safe Stain (Giotto Biotech). Western blots (WB) were performed using nitrocellulose membranes for gel transfer and goat anti-human IgA alpha chain (HRP) (Abcam) with SuperSignal™ West Pico Chemiluminescent Substrate (ThermoFischer Scientific) for antibody detection, in Trans-Blot Turbo Transfer System (Bio-Rad Laboratories, Inc).

### Production of 293T cells stably expressing human ACE2 receptor

Production of 293T cells stably expressing human ACE2 receptor was done as previously described ^43^. Briefly, VSV-G pseudotyped lentiviruses encoding human ACE2, 293ET cells were transfected with pVSV-G, psPAX2 and pLEX-ACE2 using jetPRIME (Polyplus), according to manufacturer’s instructions.

Lentiviral particles in the supernatant were collected after 3 days and were used to transduce 293T cells. Three days after transduction, puromycin (Merck, 540411) was added to the medium, to a final concentration of 2.5 μg/ml, to select for infected cells. Puromycin selection was maintained until all cells in the control plate died and then reduced to half. The 293T-Ace2 cell line was passaged six times before use and kept in culture medium supplemented with 1.25 μg/ml puromycin.

### Production and titration of spike pseudotyped lentiviral particles

To generate spike pseudotyped lentiviral particles, 6×10^6^ 293ET cells were co-transfected with 8.89ug pLex-GFP reporter, 6.67μg psPAX2, and 4.44μg pCAGGS-SARS-CoV-2-S_trunc_ D614G, using jetPRIME according to manufacturer’s instructions. The virus-containing supernatant was collected after 3 days, concentrated 10 to 20-fold using Lenti-XTM Concentrator (Takara, 631231), aliquoted and stored at -80°C. Pseudovirus stocks were titrated by serial dilution and transduction of 293T-Ace2 cells. At 24h post transduction, the percentage of GFP positive cells was determined by flow cytometry, and the number of transduction units per mL was calculated.

### Neutralization assay

Heat-inactivated skim breast milk and plasma samples were four-fold serially diluted and then incubated with spike pseudotyped lentiviral particles for 1h at 37°C. The mix was added to a pre-seeded plate of 293T-Ace2 cells, with a final MOI of 0.2. At 48h post-transduction, the fluorescent signal was measured using the GloMax Explorer System (Promega). The relative fluorescence units were normalized to those derived from the virus control wells (cells infected in the absence of plasma or skim breast milk), after subtraction of the background in the control groups with cells only.

### Statistical analysis

The half-maximal neutralization titre (NT_50_), defined as the reciprocal of the dilution at which infection was decreased by 50%, was determined using four-parameter nonlinear regression (least squares regression without weighting; constraints: bottom=0) (GraphPad Prism 9). The nonparametric Wilcoxon test (paired) and Man-Whitney test (unpaired) and the parametric *t* test (paired and unpaired) were used as described in figure legends. Spearman and Pearson correlation test were used in correlation analysis. ANOVA with post-hoc Turkey’s multiple comparison test and Friedman with post-hoc Dunn’s multiple comparison test were used to compare means between groups.

---

**Table S1.** Demographic data of nursing health care workers.

| **Nursing** | **Feeding duration (months)** | **Days post 1<sup>st</sup> dose** | **Days post 2<sup>nd</sup> dose** | **COVID-19 diagnostic** |
| --- | --- | --- | --- | --- |
| 1 | 16 | 10 | 10 | No |
| 2 | 12 | 10 | 11 | No |
| 3 | 7 | 8 | 7 | No |
| 4 | 13 | 13 | 13 | No |
| 5 | 21 | 13 | 10 | No |
| 6 | 23 | 8 | 10 | No |
| 7 | 13 | 8 | 8 | No |
| 8 | 11 | 16 | 9 | No |
| 9 | 4 | 10 | 10 | No |
| 10 | 15 | 12 | 12 | No |
| 11 | 9 | 9 | 7 | No |
| 12 | 13 | 8 | 9 | No |
| 13 | 3 | 8 | 9 | No |
| 14 | 13 | 7 | 10 | No |

**Table S2.** Demographic data of controls health care workers.

| **Controls** | **Days post 2<sup>nd</sup> dose** | **COVID-19 diagnostic** |
| --- | --- | --- |
| 1 | 10 | No |
| 2 | 16 | No |
| 3 | 10 | No |
| 4 | 10 | No |
| 5 | 10 | No |
| 6 | 10 | No |
| 7 | 10 | No |
| 8 | 11 | No |
| 9 | 11 | No |
| 10* | 11 | No |

\*Excluded from the study due to post-menopausal status.

## Notes

### Competing Interest Statement

The authors have declared no competing interest.

### Funding Statement

This work was supported by ESCMID and by Gilead Génese (PGG/009/2017) grants to HS, RESEARCH4COVID 19 (Ref 580) to MJA, EU H2020 projects No. 823780 and 871037 (iNEXT-Discovery) to MA, and by Fundação para a Ciência e Tecnologia (FCT) through Project MOSTMICRO-ITQB with refs UIDB/04612/2020 and UIDP/04612/2020 to MA and DA. JG, DA, MJA and HS are supported by Fundação para a Ciência e Tecnologia (FCT) through PD/BD/128343/2017, BD/147987/2019, CEECIND/02373/2020 and CEECIND/2020/01049, respectively.

### Author Declarations

This study has been approved by NOVA Medical School ethics committee, in accordance with the provisions of the Declaration of Helsinki and the Good Clinical Practice guidelines of the International Conference on Harmonization.

